# Accurate inference methods based on the estimating equation theory for the modified Poisson and least-squares regressions

**DOI:** 10.1101/2025.01.10.25320320

**Authors:** Hisashi Noma, Masahiko Gosho

## Abstract

**Objectives:** In clinical and epidemiological studies, the modified Poisson and least-squares regression analyses for binary outcomes have been used as standard multivariate analysis methods to provide risk ratio and risk difference estimates. However, their ordinary Wald-type confidence intervals can suffer from finite-sample biases in the robust variance estimators, and the coverage probabilities of true effect measures are substantially below the nominal level (usually 95%). To address this issue, new accurate inference methods are needed.

**Methods:** We propose two accurate inference methods based on the estimating equation theory for these regression models. A remarkable advantage of these regression models is that the correct models to be estimated are known, that is, conventional binomial regression models with log and identity links. Using this modeling information, we first derive the quasi-score statistics, whose robust variances are estimated using the correct model information, and then propose a confidence interval based on the regression coefficient test using *χ*^2^ -approximation. To further improve the large sample approximation, we propose adapting a parametric bootstrap method to estimate the sample distribution of the quasi-score statistics using the correct model information. In addition, we developed an R package, `rqlm` (https://doi.org/10.32614/CRAN.package.rqlm), that can implement the new methods via simple commands.

**Results:** In extensive simulation studies, the coverage probabilities of the two new methods clearly outperformed the ordinary Wald-type confidence interval when the regression function assumptions were correctly specified, especially in small and moderate sample settings. We also illustrated the proposed methods by applying them to an epidemiological study of epilepsy. The proposed methods provided wider confidence intervals, reflecting statistical uncertainty.

**Conclusions:** The current standard Wald-type confidence intervals may provide misleading evidence. Erroneous evidence can potentially influence clinical practice, public health, and policymaking. These possibly inaccurate results should be circumvented using effective statistical methods. These new inference methods would provide more accurate evidence for future medical studies.

## Introduction

Logistic regression has been a standard multivariate analysis method for analyzing binary outcome data in clinical and epidemiological studies. However, the odds ratio is difficult to interpret as an effect measure. While the causal odds ratio can be formally defined using a counterfactual framework, it can only be interpreted as an approximation of the risk ratio when the frequency of events is small [1, 2]. Thus, the use of risk ratios and risk differences is recommended as an alternative in various guidelines; for example, the CONSORT statements recommend reporting relative and absolute measures of effect when reporting the results of clinical trials [3, 4].

Owing to the substantial limitations of logistic regression, other binomial regression models have conventionally been considered using the log or identity link functions to provide risk ratio and risk difference estimators [5]. However, the values of these binomial regression models are not limited within the range [0, 1], and the maximum likelihood (ML) estimates often cannot be defined in practice [6, 7]. To address these issues, Zou [8] and Cheung [9] proposed modified Poisson and least-squares (Gaussian) regression analyses, which provide consistent risk ratio and risk difference estimators without computational difficulties. Their ideas were to formally fit the Poisson and least -squares regression models to the binary outcome data and calculate the regression coefficient estimates by the framework of the generalized linear model (GLM) [10]. The resultant estimators then provide consistent estimates of risk ratios and risk differences based on the estimating equation theory of the GLM [11, 12], even if the distributional assumptions are misspecified. In addition, the variance estimators should be changed to the sandwich variance estimators [13].

One relevant issue in logistic regression is the serious bias of the regression coefficient estimator for small samples, and various correction methods have been discussed [14, 15]. Recently, Uno et al. [16] have shown that the same bias can occur in the modified Poisson regression analysis, although this phenomenon does not occur in the modified least-squares regression. More importantly, they showed that the robust variance estimators for both regression analysis methods can be biased and that the resultant Wald-type confidence intervals can seriously underestimate the actual statistical errors in small or moderate sample settings. These properties can lead to misleading evidence in clinical and epidemiological studies; therefore, accurate alternative statistical inference methods are needed.

In this study, we propose new confidence intervals for the modified Poisson and least-squares regression analyses based on the estimating equation theory, especially for accurate inferences in small or moderate sample settings. A remarkable advantage of these regression models is that we know the correct models to be estimated, that is, conventional binomial regression models with log and identity links. Using this modeling information, we first derive the quasi-score statistics for these regression models, whose robust variances are estimated using the correct model information. Quasi-score-based inferences have been discussed for various pseudo-likelihood inferences (e.g., Mantel– Haenszel methods [17, 18]) and are known to have favorable properties compared with naïve Wald-type inferences. We subsequently propose a confidence interval based on the quasi-score test using *χ*^2^ -approximation. To further improve small sample approximations, we propose adapting a parametric bootstrap method to estimate the sample distribution of the quasi-score statistics using the correct model information. Through extensive simulation studies, we showed that the coverage probabilities of the two new methods clearly outperform the ordinary Wald-type confidence interval. We also illustrate the proposed methods through their application in an epidemiological study of epilepsy. We developed an R package, `rqlm` (https://doi.org/10.32614/CRAN.package.rqlm), that can implement the new methods via simple commands.

### Modified Poisson and least-squares regressions

We consider a cohort study consisting of *n* participants with binary outcomes *Y*_1_, …, *Y*_*n*_ (= 1: event occurred, = 0: event did not occur) and covariates ***x***_*i*_ = (*x*_*i*1_, *x*_*i*2_, …, *x*_*ip*_)^*T*^ for the *i*th subject (*i* = 1, …, *n*). Conventionally, binomial regressions with log link and identity link functions have been considered for multivariate analyses of risk ratios and risk differences; however, they involve serious theoretical difficulties in defining the ML estimates because the values of the regression functions do not fall within [0, 1] [6, 7]. Zou [8] and Cheung [9] proposed the modified Poisson and least-squares regressions as effective methods for multivariate analyses. Their ideas involved formally fitting the Poisson and least-squares regression models to the binary outcome data.

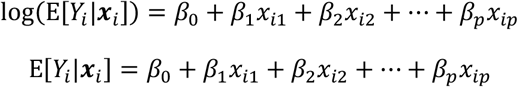

The resultant quasi-ML estimators 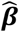 of the regression coefficients ***β*** = (*β*_0_, *β*_1_, …, *β*_*p*_)^*T*^ become consistent estimators of the log-transformed risk ratios and risk differences on the target population [16]. The principle of these estimating methods is based on the estimating equation theory of the GLM [12]; that is, the estimating functions are unbiased even if the distribution forms are misspecified as long as the functional forms of the regression functions are correctly specified. In particular, for the modified least-squares regression, the quasi-ML estimator becomes a linear unbiased estimator [16]; 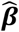 is an unbiased estimator for the regression coefficients of the binomial regression function [16]. The standard errors of 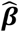 of both models are consistently estimated by the sandwich variance estimator [13].

### Confidence intervals for risk ratios and risk differences

#### Confidence intervals based on the quasi-score statistics

The modified Poisson and least-squares regressions are effective methods for multivariate analyses of risk ratios and risk differences; however, their ordinary Wald-type confidence intervals can seriously underestimate statistical errors in small or moderate sample settings [16]. To address these issues, we first derive quasi-score tests for the regression coefficients. The two models are formulated as specific cases of the GLM, and the quasi-likelihood estimating functions are expressed as

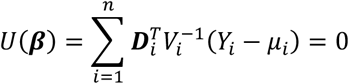

where *μ*_*i*_ is the mean function (= exp (***β***^*T*^***x***_*i*_) for the Poisson model and = ***β***^*T*^***x***_*i*_ for the Gaussian model and ***D***_*i*_ = *∂μ*_*i*_/*∂* ***β***; moreover, *V*_*i*_ = *ν*(*μ*_*i*_) is the variance function of the outcome variable (= *μ*_*i*_ for the Poisson model and = 1 for the Gaussian model) (*i* = 1, …, *n*). If the variance functions are correctly specified, the covariance matrices of *U*(***β***) become the Fisher information matrices *I*(***β***) = −*E*[*∂U*(***β***)/*∂****β***]. However, the variance functions are misspecified for these cases; therefore, they become the robust covariance matrices, *J*(***β***) = *E*[*U*(***β***)*U*^*T*^(***β***)]. In addition, a special feature of these inferences is that the correct models are known to us and expectations can be substituted into the binomial regression models. Then, the concrete forms of the models are expressed as

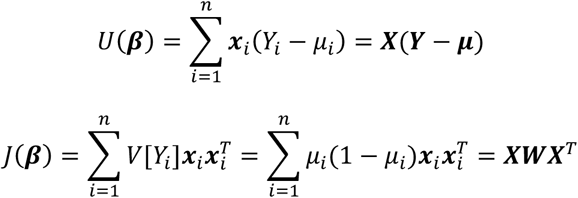

where ***Y*** = (*Y*_1_, …, *Y*_*n*_)^*T*^, ***μ*** = (*μ*_1_, …, *μ*_*n*_)^*T*^, ***X*** = (***x***_1_, …, ***x***_*n*_), and ***W*** = diag{*μ*_1_(1 − *μ*_1_), …, *μ*_*n*_(1 − *μ*_*n*_)}. Although the definitions of the mean function *μ*_*i*_ differ between the two models, the function forms are the same. Note that *μ*_1_, …, *μ*_*n*_ should be truncated on [0, 1] on ***W*** because the individual variance functions substantially estimate the variances of the binomial variables and should not be negative values. However, those in *U*(***β***) should not be truncated; if they are truncated, the quasi-score functions are biased and unrealistically singular results can be obtained. Furthermore, we note that *E*[*U*(***β***)] = **0** and *V*[*U*(***β***)] = *J*(***β***) without large sample approximations. The quasi-score test statistics for the joint null hypotheses H0: ***β*** = ***β***_null_ are constructed using the exact means and covariance matrices. However, these null hypotheses are usually outside the scope of interest in practice.

We consider tests for composite null hypotheses H0: *β*_1_ = *β*_1,null_ that correspond to the hypothesis tests for individual risk ratios and risk differences; without loss of generality, we consider the tests of regression coefficients of first explanatory variables and denote ***β*** = (*β, β*, …, *β*) _*T*_. The quasi-score statistics are then constructed as follows:

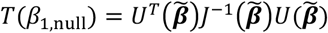

where 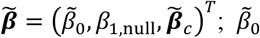 and 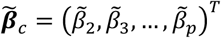 are the constrained quasi-ML estimates of {*β*_0_, ***β***_*c*_} under H0. The constrained quasi-ML estimates can be calculated via the same modified Poisson and least-squares regression analyses when *x*_1*i*_ is dropped from the explanatory variables and offsets *β*_1,null_*x*_1*i*_ are added (*i* = 1, …, *n*). Under the null hypotheses, the quasi-score test statistics follow the *χ*^2^-distribution with approximately one degree of freedom [19]. Moreover, using these quasi-score tests, we can construct the 100 × (1 − *α*)% confidence intervals of *β*_1_ using the sets of null values that satisfy

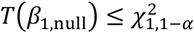

where 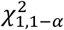 is the upper *α*th percentile of the *χ*^2^ -distribution with one degree of freedom. The confidence limits can be calculated using adequate numerical methods (e.g., the bisectional methods [20]).

As shown in the simulation studies, confidence intervals based on the quasi-score statistics generally have favorable properties compared with the ordinary Wald-type confidence intervals obtained using the standard sandwich variance estimators. However, confidence intervals when using the large sample *χ*^2^ approximations still have limitations with respect to achieving sufficient coverage performance in small sample settings [21]. In addition, more accurate approximations of the sample distributions are needed for valid inferences.

#### Bootstrap confidence intervals based on the quasi-score statistics

To improve the accuracy of approximations of the sample distributions of the quasi-score statistics, we propose using the bootstrap method. Again, we focus on the advantage of these regression methods—that the correct distributional assumptions for the target population are known (binomial regression models). Therefore, we propose performing parametric bootstrap resampling from the “correct” binomial regression models by substituting the regression coefficients ***β*** for the null value and constrained quasi-ML estimates. The bootstrap algorithm for the tests of H0: *β*_1_ = *β*_1,null_ is as follows.

#### Algorithm (bootstrap tests for the quasi-score statistics)

1. For the modified Poisson and least-squares regression models, compute the constrained quasi-ML estimates 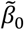 and 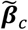 under H0: *β*_1_ = *β*_1,null_.
2. Resample 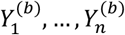 from the binomial regression models with log or identity links whose regression coefficients ***β*** are fixed to 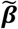

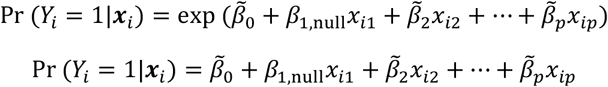

via parametric bootstraps, *B* times (*i* = 1, …, *n*; *b* = 1,2, …, *B*). Note that, if the values of the regression functions on the right-hand sides of these equations exceed the range [0, 1], then they should be truncated at 0 or 1. In addition, the design matrix ***X*** is not altered from the original data across resampling.
3. Compute the quasi-score statistic *T*^(*b*)^(*β*_1,null_) for the *b* th bootstrap sample 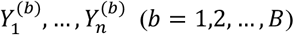.
4. Calculate the empirical distribution function of *T*^(1)^(*β*_1,null_), …, *T*^(*B*)^(*β*_1,null_) — specifically, 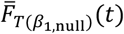 —which is the bootstrap estimate of the sample distribution of *T*(*β*_1,null_)
5. Implement the hypothesis test for *T*(*β*_1,null_) using 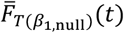 for the reference distribution.

Because accurate tail area estimation of the null distribution requires a large number of replications [22], the number of bootstrap resamplings *B* should be sufficiently large (usually at least 1000).

The corresponding 100 × (1 − *α*)% confidence intervals of *β*_1_ can be constructed using the sets of *β*_1,null_ that satisfy

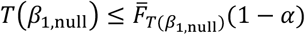

The confidence limits can also be calculated using adequate numerical methods (e.g., the bisectional methods [20]). The parametric bootstrap approach effectively uses the distributional information of the correct models; thus, the approximation is expected to improve compared with the naïve asymptotic normal approximation. The actual performances are demonstrated in simulation studies. Note that the validity of the proposed confidence intervals requires correct specifications of the regression functions, which can be violated if the pivotal assumptions are incorrect. However, the validity of other standard confidence intervals (e.g., Wald-type and nonparametric bootstrap confidence intervals) is also violated under these conditions.

#### Software

We developed an R package, `rqlm` (https://cran.r-project.org/web/packages/rqlm), to perform all of the proposed methods via simple commands. An example of the R code used to implement these methods is provided in the Supplementary Materials.

### Simulations

To illustrate the operating characteristics of the proposed methods, we conducted simulation studies. For data generation, we considered the binomial regression models with log and identity link functions, and parameter settings were selected to mimic the epidemiological study of epilepsy described in the next section. Four explanatory variables were considered: *x*_*i*1_ was the main treatment/exposure variable that followed a Bernoulli distribution with probability 0.20 or 0.10; *x*_*i*2_ was a confounding variable that followed a Bernoulli distribution with probability 0.773 and had a correlation with *x*_*i*1_ through measurement of the odds ratio [OR] Pr (*x*_*i*1_ = 1, *x*_*i*2_ = 1)Pr (*x*_*i*1_ = 0, *x*_*i*2_ = 0)/Pr (*x*_*i*1_ = 0, *x*_*i*2_ = 1)Pr (*x*_*i*1_ = 1, *x*_*i*2_ = 1) = 25, 15, and 5; *x*_*i*3_ followed a Bernoulli distribution with probability 0.455; and *x*_*i*4_ followed *N*(29.0, 7.37). The outcome variable *Y*_*i*_ was then generated from a Bernoulli distribution with probability

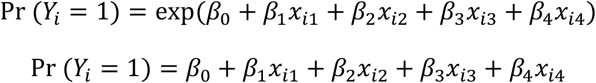

The intercept *β*_0_ was set by controlling for the overall event rate of the cohort; the event rate was varied as 0.40 and 0.20. The other regression coefficients were set as (*β*_1_, *β*_2_, *β*_3_, *β*_4_) = (0.205, −0.271, 0.000, 0.153) for the former model and = (0.116, −0.041, 0.0037,0.023) for the latter model. We considered the sample size *n* = 20, 30, …, 100; small and moderate sample settings in which the robust variance estimator may not perform well. If separation or quasi-separation situations occur, the corresponding cases are excluded from the experiments because the quasi-ML estimates might not be obtained under these settings. We performed 2,000 simulations for the 108 scenarios for both the risk ratio and risk difference regression models.

For comparisons, we analyzed the individual dataset using the ordinary modified Poisson and least-squares regressions and Wald-type confidence intervals obtained via the standard sandwich variance estimator, HC3-type bias-adjusted sandwich variance estimator [23], Firth-type correction method using the bias-adjusted sandwich variance estimator [16], nonparametric bootstrap confidence intervals [22], and Fisher’s *z*-transformation proposed by Zou and Donner [24] (only for the modified least-squares regression). We then applied the two proposed confidence intervals based on the quasi-score statistics and bootstrap approach. For the bootstrap methods, we performed 2,000 bootstrap resamplings to calculate the bootstrap distributions. We assessed the coverage probabilities of the 95% confidence intervals for *β*_1_ of the three methods. The R codes used to implement the simulations are provided in the Supplementary Materials.

The results of the simulation studies are presented in Figures 1 and 2 for the modified Poisson regression and in Figures 3 and 4 for the modified least-squares regression. The empirical coverage rates of the 95% confidence intervals for the 2,000 simulations were plotted. For the modified Poisson regression, the four conventional confidence intervals suffered from undercoverage in small and moderate sample settings. These results were due to the biases of both the regression coefficient estimates and robust standard error estimates. The improved methods based on higher-order approximations (HC3 and Firth-type correction) still suffered from invalidity in these settings. The degree of undercoverage was serious if the event rate or exposure rate became too small and the correlations of *x*_*i*1_ and *x*_*i*2_ became too large. In addition, the two proposed confidence intervals retained coverage probabilities around the nominal level (95%) for most scenarios. The quasi-score-based confidence intervals could reflect undercoverage bias in some small sample settings (generally, *n* ≤ 50). However, the coverage properties were consistently favorable compared with those of the conventional confidence interval. In addition, as expected, the proposed bootstrap confidence intervals exhibited better coverage probabilities. Even for small sample settings (*n* = 20), the coverage probabilities consistently had values around the nominal level (95%).

**Figure 1.**
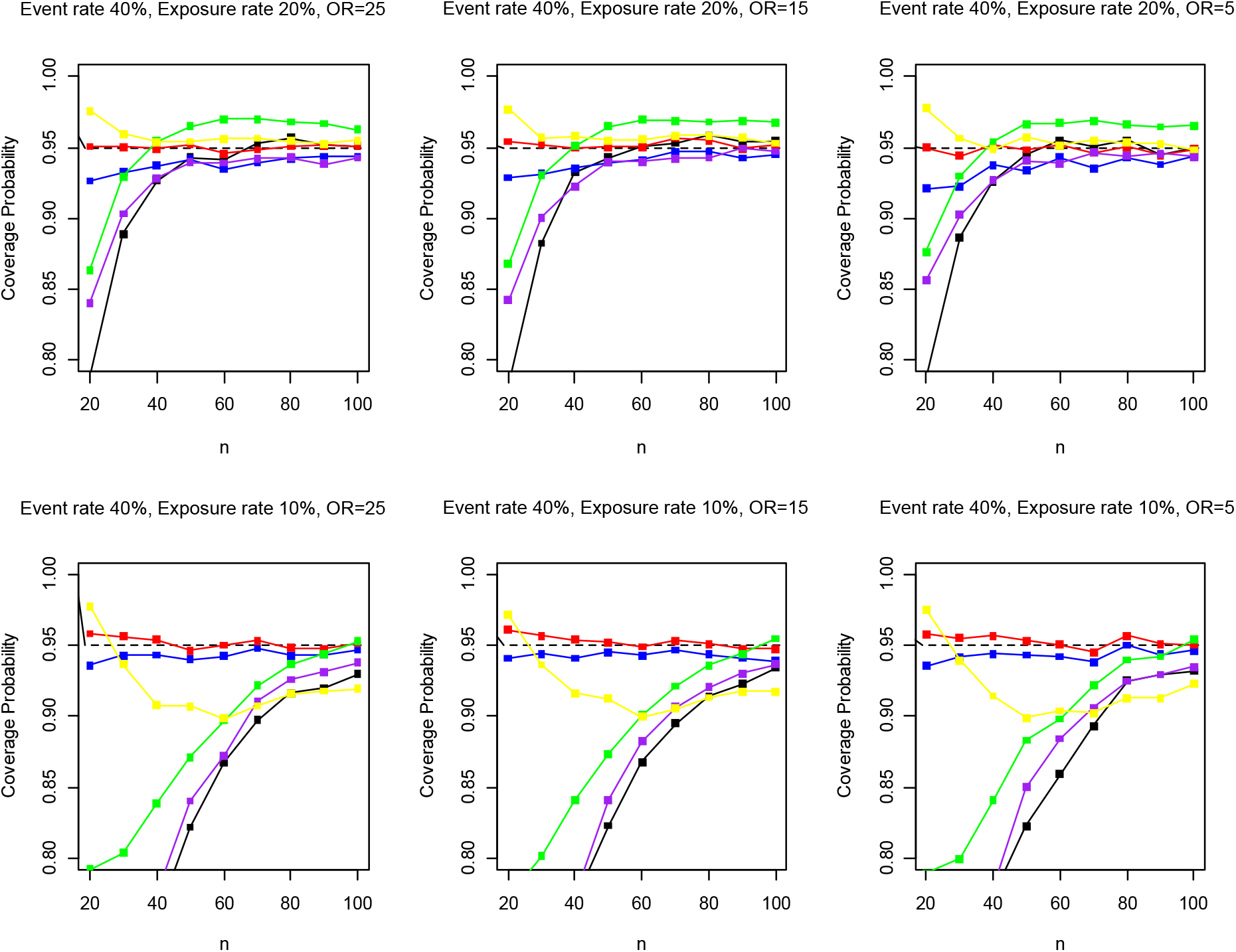
Results of simulations for the modified Poisson regression (black: ordinary Wald CI, green: Wald CI using HC3-estimator, yellow: Wald CI using Firth-type estimator, purple: nonparametric bootstrap CI, blue: quasi-score CI, red: bootstrap CI by the quasi-score statistic; CI: confidence interval).

**Figure 2.**
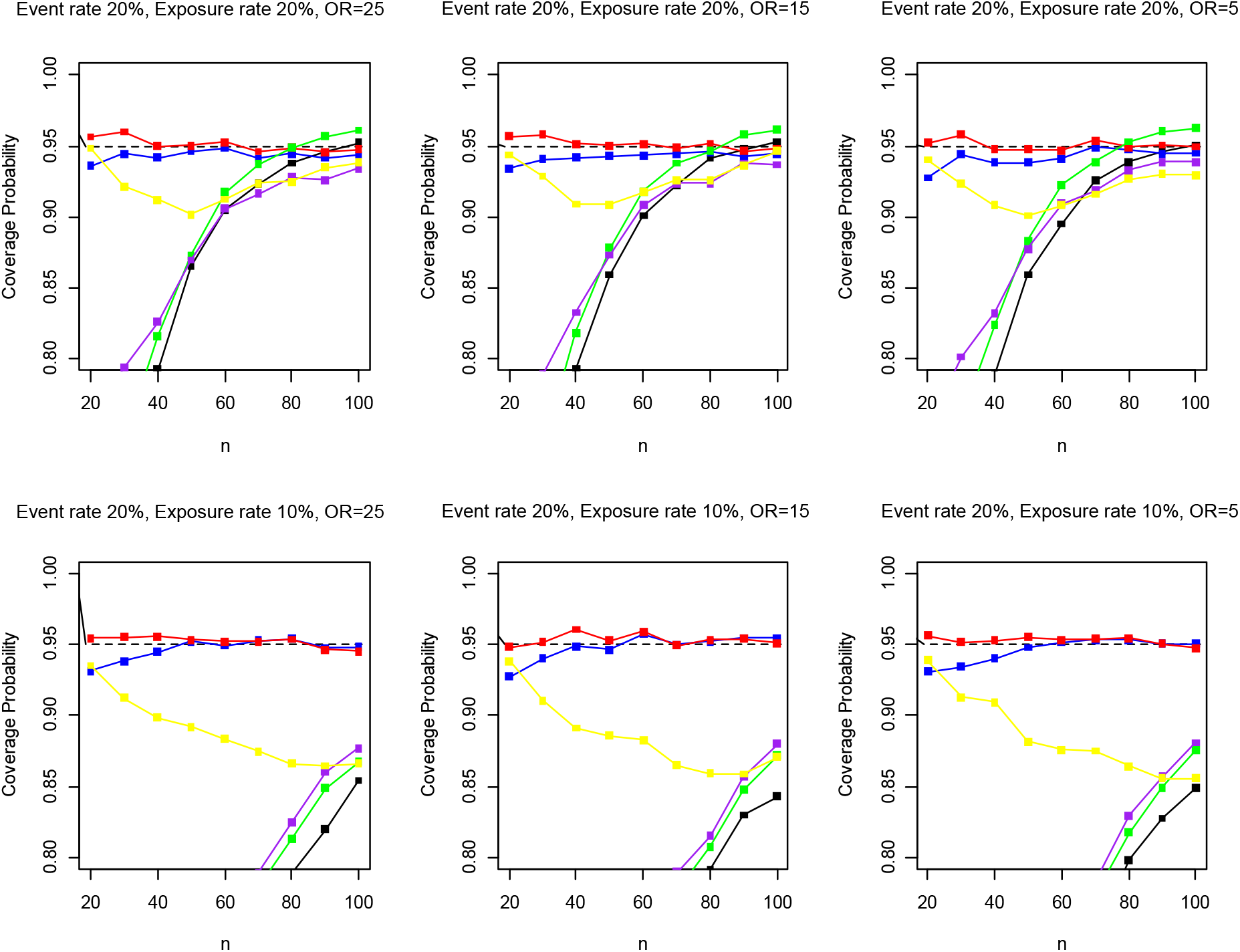
Results of simulations for the modified Poisson regression (black: ordinary Wald CI, green: Wald CI using HC3-estimator, yellow: Wald CI using Firth-type estimator, purple: nonparametric bootstrap CI, blue: quasi-score CI, red: bootstrap CI by the quasi-score statistic; CI: confidence interval).

**Figure 3.**
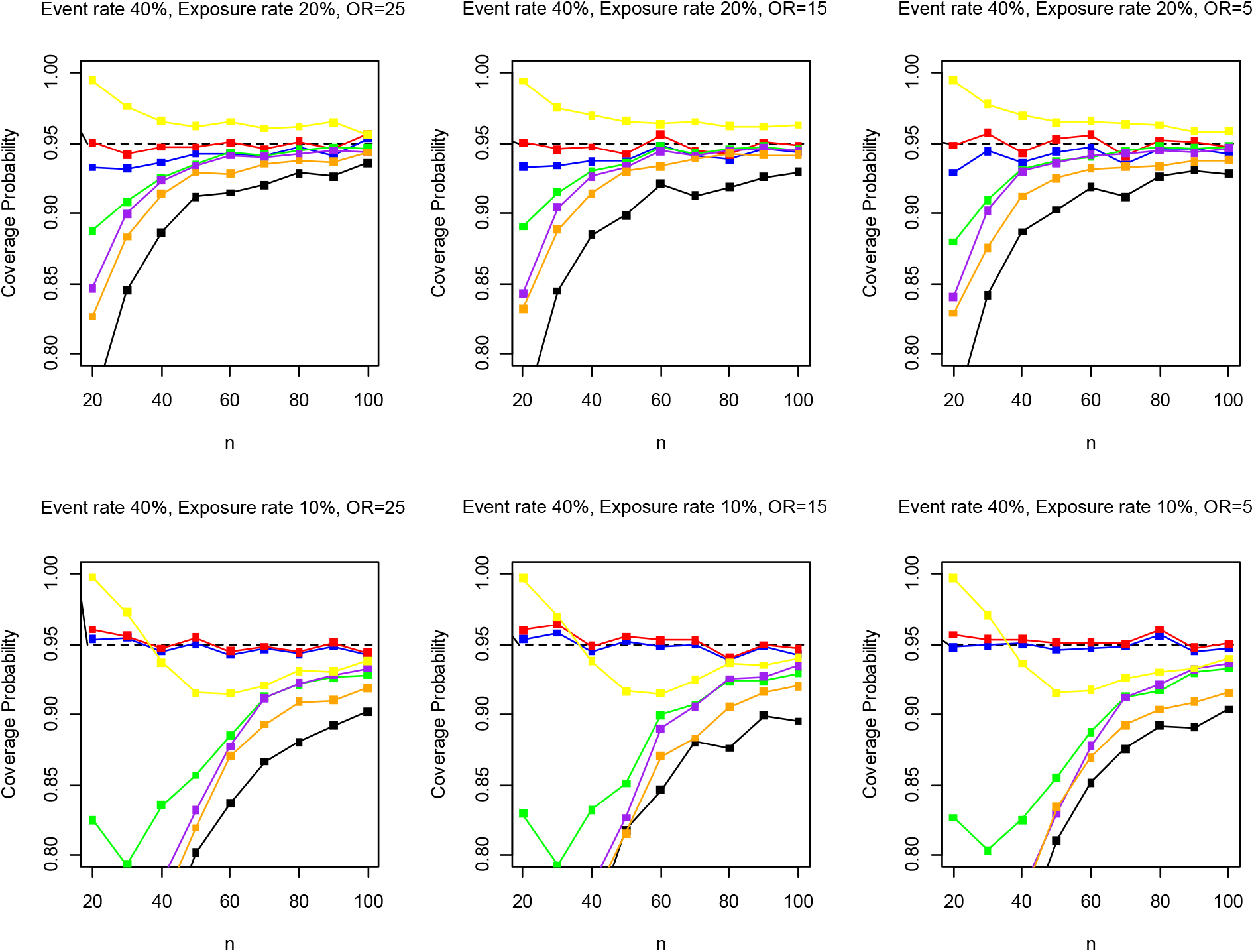
Results of simulations for the modified least-squares regression (black: ordinary Wald CI, green: Wald CI using HC3-estimator, yellow: Wald CI using Firth-type estimator, purple: nonparametric bootstrap CI, orange: Fisher’s z-transformation, blue: quasi-score CI, red: bootstrap CI by the quasi-score statistic; CI: confidence interval).

**Figure 4.**
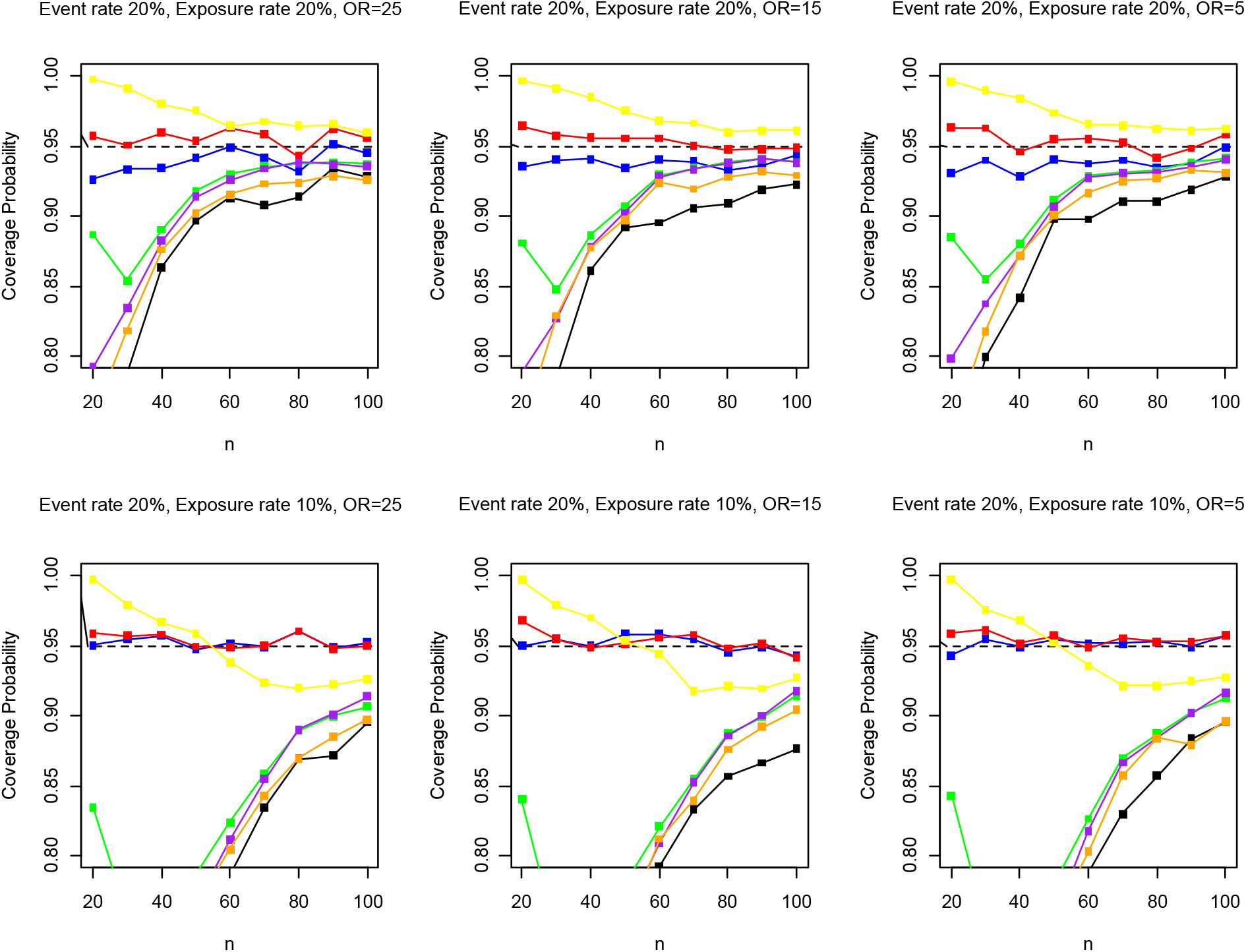
Results of simulations for the modified least-squares regression (black: ordinary Wald CI, green: Wald CI using HC3-estimator, yellow: Wald CI using Firth-type estimator, purple: nonparametric bootstrap CI, orange: Fisher’s z-transformation, blue: quasi-score CI, red: bootstrap CI by the quasi-score statistic; CI: confidence interval).

For the modified least-squares regression, the overall simulation results were similar to those of the modified Poisson regression simulations. The conventional confidence intervals were generally prone to undercoverage in small and moderate samples. The quasi-ML estimates corresponded to a linear unbiased estimator [16]; thus, there were no biases in the point estimator. However, robust variance estimators were seriously biased in small and moderate sample settings. Moreover, the degree of undercoverage became serious if the event rate or exposure rate became excessively small and the correlations of *x*_*i*1_ and *x*_*i*2_ became large. In addition, the proposed confidence intervals exhibited favorable coverage properties. The quasi-score-based confidence interval was prone to undercoverage in some settings; however, the coverage properties were much better than those of the conventional methods. Additionally, the proposed bootstrap confidence interval consistently improved the coverage properties. Even in small sample settings (*n* = 20), the coverage probabilities consistently had values close to the nominal level (95%). These simulation results indicate that the proposed bootstrap confidence interval provided accurate interval estimates and that validity was retained even in small and sparse data settings.

### Applications

To illustrate the usefulness of the proposed methods, we applied them to an epidemiological study of epilepsy conducted by Arai et al. [25], which was a retrospective cohort study that evaluated the factors associated with the employment status of patients with a history of childhood-onset drug-resistant epilepsy (*N* = 56). We analyzed cohort data using the modified Poisson and least-squares regressions. The outcome was employment status (1 = non-employment, 0 = employment; the number of events was 14), and four explanatory variables were included: age at follow-up (`age`; continuous), gender (`gender`; male or female), mood disorder symptoms (`symptoms`; yes or no), and graduating from a school for special needs education (`education`; yes or no). The last two variables were highly correlated and were strongly associated with the outcome through univariate analyses [25].

Table 1 presents the results of the study. In many cases, the confidence intervals based on the quasi-score statistics and bootstrap approach were asymmetric around the quasi-ML estimates. In some cases, the locations of the confidence intervals differed substantially from those of the Wald-type confidence intervals. This phenomenon is known to occur in efficient score-based confidence intervals [21]. For the modified Poisson regression, only the 95% Wald-type confidence interval for `education` did not cover the null value (= 1). The two proposed confidence intervals also did not cover 1; however, the bootstrap confidence interval was relatively narrow compared with the Wald-type confidence interval. For `symptoms`, the confidence interval based on the quasi-score statistic was narrower than the Wald-type confidence interval and did not cover 1; however, the bootstrap confidence interval was much wider and did not cover 1. These results might be influenced by the strong correlation between these two covariates. In addition, for the modified least-squares regression, the Wald-type confidence interval of `symptoms` was substantially widened using the bootstrap-based approach. Additionally, the bootstrap confidence interval for `education` was wider than the Wald-type confidence interval. Considering the operating characteristics shown by the simulations, the conclusions obtained from the ordinary Wald-type confidence intervals might be misleading, and improved methods would likely provide more precise evidence.

**Table 1.** Results of the modified Poisson and least-squares regression analyses for the epilepsy epidemiological study (*N* = 56) ^†^.

|  | Risk ratio estimation using the modified Poisson regression |  |  | Risk difference estimation using the modified least-squares regression |  |  |
| --- | --- | --- | --- | --- | --- | --- |
|  | Quasi-ML estimate<br>(Wald 95%CI) | 95%CI by the quasi-score statistic | Bootstrap 95%CI by<br>the quasi-score<br>statistic | Quasi-ML estimate<br>(Wald 95%CI) | 95%CI by the quasi-score statistic | Bootstrap 95%CI by<br>the quasi-score<br>statistic |
| Age at follow-up | 1.013<br>(0.982, 1.045) | (0.981, 1.050) | (0.978, 1.067) | 0.006<br>(−0.003, 0.014) | (−0.003, 0.014) | (−0.003, 0.022) |
| Gender (male vs. female) | 1.372<br>(0.638, 2.950) | (0.669, 3.030) | (0.614, 3.509) | 0.097<br>(−0.089, 0.283) | (−0.063, 0.237) | (−0.071, 0.426) |
| Mood disorder symptoms<br>(yes vs. no) | 2.076<br>(0.781, 5.519) | (1.001, 3.230) | (0.882, 7.385) | 0.498<br>(0.153, 0.842) | (0.103, 0.721) | (0.067, 1.000) |
| Graduating from a school<br>for special needs education<br>(yes vs. no) | 0.240<br>(0.075, 0.765) | (0.100, 0.686) | (0.028, 0.721) | −0.422<br>(−0.758, −0.087) | (−0.688, −0.098) | (−0.781, −0.086) |

<sup>†</sup> CI: confidence interval.

## Discussion

The modified Poisson and least-squares regressions have been widely used in recent clinical and epidemiological studies because they provide interpretable effect measure estimates without computational difficulties. Considering the difficulty in interpreting the odds ratios, these methods will be increasingly adopted in future studies as effective alternatives to the conventional logistic regression. This study demonstrates how ordinary Wald-type confidence intervals in these settings are prone to serious undercoverage with small or moderate sample sizes. As clearly shown in the real data example, the ordinary confidence intervals might provide misleading evidence, which can potentially influence clinical practice, public health, and policymaking. These possibly inaccurate results should be circumvented using effective statistical methods. The new accurate confidence intervals and numerical evidence provided in this study will be useful for future medical studies.

In the simulation-based evidence, the ordinary Wald-type and other conventional confidence intervals showed serious undercoverage performance. For the modified Poisson regression, a relevant reason for the invalid property is the small sample bias; although bias of the ML estimator for the GLM is well known [26], a similar bias can occur for the quasi-ML estimator for misspecified models. Effective solutions to address this bias are the Firth-type bias correction [16] and higher-order bias correction methods [26]. In addition, for the modified least-squares regression, the resultant estimator is unbiased because it corresponds to a linear unbiased estimator, and no corrections are required if this criterion is considered [16]. However, the undercoverage properties shown in the simulation studies are severe, and bias of the ordinary robust variance estimator should be adequately addressed in practice. The proposed methods are expected to provide an effective solution to this problem.

A fundamental assumption to ensure consistency of the quasi-ML estimators is that the regression models are correctly specified, similar to other regression models involving ordinary logistic regression analysis. In addition, the current available inference methods employed in the simulation studies assume that the regression models are correctly specified. This is a practically unverifiable assumption but should be carefully considered in practice. In particular, misspecifications of the regression models can violate the validity of the proposed confidence intervals, and the validity of the variance estimators is violated if the regression models are misspecified. Similar problems can occur for other current standard confidence intervals (e.g., Wald-type and nonparametric bootstrap confidence intervals), and further developments of alternative robust methods to address the model misspecification problem are relevant issues in future studies. An effective approach to resolve this issue is to adopt nonlinear regression models (e.g., spline models), especially in cases where continuous covariates are modelled [27]. In addition, other regression analysis methods have been proposed to estimate interpretable effect measures [28-31], and similar discussions are beneficial for these alternative effective approaches. Developments of new methods for these alternative regression methods would also be relevant future issues.

Another future issue is adapting improved robust variance estimators for the Wald-type confidence interval [32]. Although the accuracy of the existing improved robust variance estimators is better than that of the ordinary sandwich variance estimator, deterministic conclusions cannot be provided for their relative performances [32] because all methods are based on approximations (e.g., higher-order approximations). Although simulation-based numerical evidence can provide good case-by-case comparative performance, they would not be generic properties. However, the bootstrap-based approach proposed in the present study is based on the quasi-score statistic, which effectively uses information from the null hypothesis and adapts the flexible sample distribution estimate. Therefore, we believe that it is one of the most effective methods among the competing methods. The proposed methods can be used as accurate and effective alternatives to the ordinary Wald-type inference methods in clinical and epidemiological studies.

## Acknowledgements

The authors would like to thank Dr. Y. Arai (Tottori University) for permission to use the valuable data and K. Nakazono (The Institute of Statistical Mathematics) for his helpful comments on the earlier draft.

## Research Funding

This study was supported by Grants-in-Aid for Scientific Research from the Japan Society for the Promotion of Science (grant numbers: JP23K11931, JP22H03554, JP24K21306, and JP23H03063).

## Data availability

R package for implementing the proposed methods is available at CRAN (https://cran.r-project.org/web/packages/rqlm).

